# RMNCH Service Availability and Readiness of Restructured Primary Health-care Facilities in Ethiopia: a multi-region cross-sectional assessment

**DOI:** 10.64898/2026.09.26.26364072

**Authors:** Mesele Damte Argaw, Geteneh Moges, Agumasie Semahegn, Gizachew Tadele Tiruneh, Rediet Daniel, Mikiyas Teferi, Shegaw Mulu Tarekegn, Derebe Tadesse Abate, Sisay Menji, Yayeh Kassa, Firew Solomon, Aklog Getnet, Wondesen Nigatu, Addis Girma, Mebrie Belete, Yenegeta Walelign, Berhanu Tamiru Tirfie, Abdulhelim Tuffa, Muluken Dessalegn Muluneh, Lidiya Tefera, Frank DelPizzo, Kidist Lulu Hagos, Temesgen Ayehu, Israel Ataro Otoro, Helina Worku, Addis Tamire, Desalew Emaway, Lisanu Taddesse, Misrak Makonnen, Thandisizwe Redford Mavundla, Brittany Hagedorn

## Abstract

**Background:** A responsive and resilient health system that is informed by community needs and guided by evidence-based interventions is fundamental to delivering quality essential health services and achieving Universal Health Coverage (UHC) and the Sustainable Development Goals (SDGs). Ethiopia committed to improve access to and quality of essential health services through the Health Extension Program Optimization (HEPO) roadmap, which restructured primary healthcare facilities into three categories i.e., Basic Health Posts (BHPs), Comprehensive Health Posts (CHPs), and Health Extension Program units imbedded within Health Centers (HCs). However, evidence on the reproductive, maternal, newborn, and child health (RMNCH) service availability and readiness of these restructured primary health-care facilities remains limited.

**Methods:** This multi-region cross-sectional facility based study was conducted between December 2024 and February 2025. A structured questionnaire adapted from the World Health Organization Service Availability and Readiness Assessment (SARA) was used to measure service availability, general facility readiness, and RMNCH service-specific readiness using predefined domains and tracer items. Differences across facility types were assessed using Kruskal-Wallis or categorical association tests, with Benjamini-Hochberg adjustment for multiple comparisons. Multivariable linear regression estimated the adjusted differences in overall RMNCH readiness, controlling for region, urban-rural residence, and log-transformed catchment population, with robust standard errors clustered at woreda level.

**Results:** A total of 305 primary health care facilities were assessed, including 107 BHPs (35.1%), 142 CHPs (46.6%), and 56 HCs (18.4%). Nearly all BHPs were located in rural settings (99.1%), compared with 86.6% of CHPs and 71.4% of HCs. The median general health facility readiness were increased significantly across facility levels, from 33.1% (IQR: 24.4-40.7) in BHPs to 62.9% (IQR: 54.1-71.4) in CHPs and 89.6% (IQR: 81.3-93.6) in HCs (p<0.001). And the median overall RMNCH service readiness was 67.9%, 77.8%, and 91.1% for BHPs, CHPs, and HCs, respectively (p<0.001). skilled Birth Attendance readiness was demonstrated the largest gap between facility types. Compared with CHPs, BHPs scored 9.9 percentage points lower in adjusted RMNCH service readiness (95% CI: -14.7 to -5.1; p<0.001), while HCs scored 8.7 points higher (95% CI: 1.7-15.8; p=0.015).

**Conclusions:** The restructured primary healthcare facilities expanded access to essential services, particularly through CHPs, which occupy an intermediate position between BHPs and HCs through substantial expansion of RMNCH service packages for underserved communities. Nevertheless, significant gaps remain in staffing, infrastructure, commodities, diagnostics, and childbirth readiness. Future HEPO scale-up should prioritize strengthening childbirth-readiness packages and ensuring reliable supply chains for essential medicines and diagnostics.

## Introduction

A responsive and resilient health system, informed by community engagement, is essential for delivering quality essential health services. Such a system, guided by evidence-based interventions, plays a critical role in achieving Universal Health Coverage (UHC) and the Sustainable Development Goals (SDGs) [1]. RMNCH services are central to primary health care and play a critical role in strengthening health systems. They contribute significantly to improving population health outcomes and advancing progress toward UHC [2,3]. Although globally maternal and under-five mortality declined substantially between 2000 and 2020, sub-Saharan Africa continues to carry a disproportionate share of preventable deaths [4,5]. These outcomes are patterned by socioeconomic position, geography, gender and unequal access to effective services [6–9].

The World Health Organization (WHO) health systems framework identifies six interdependent building blocks that are essential for health system performance: service delivery, health workforce, health information systems, access to essential medicines and technologies, health financing, and leadership and governance. Together, these components provide a comprehensive foundation for strengthening health systems and improving population health outcomes [10,11]. Ethiopia has made substantial investments in strengthening its community-based primary health care system through the Health Extension Program (HEP), which was introduced in 2003. The program has expanded access to essential health services through a network of more than 17,587 health posts and approximately 47,540 Health Extension Workers (HEWs) deployed across the country [12,13]. Health posts have improved physical access to health promotion, disease prevention and selected curative services, particularly in rural areas [14]. Nevertheless, limitations in infrastructure, staffing, commodities, referral systems, epidemiological shifts and service quality continue to constrain maternal and newborn outcomes [15,16].

To address limitations of the original HEP, the Ministry of Health endorsed the 15-year Health Extension Program Optimization (HEPO) Roadmap (2020-2035). The roadmap was developed in response to epidemiological and demographic transitions, as well as evolving community expectations, with a strong emphasis on improving quality, equity, accountability, and responsiveness within the health system [17–19]. It differentiates service packages according to settlement and livelihood context. Health posts were mapped in relation to their supervising health centers and categorized as BHPs, CHPs or HEP units embedded within HCs according to accessibility and local service needs [17,18,20]. Expansion of facility mandates consequently requires corresponding increases in the number and professional mix of health workers, infrastructure, equipment and commodities.

Studies in Kenya, Nigeria, Nepal and Bangladesh have documented substantial variation in primary-care service availability and readiness [21–24]. Ethiopian evidence has generally examined health posts, health centers or specific services separately [20,25–29]. Direct comparisons of BHPs, CHPs, and HCs are essential to determine whether the restructured primary healthcare facility has established a coherent gradient of service readiness and capacity, and to identify areas requiring additional investment and support. Therefore, this assessment aimed to compare: (1) general facility readiness; (2) service-specific readiness for ANC, SBA, and under-five child health services; (3) the availability of RMNCH services; and (4) facility-level factors associated with overall readiness across BHPs, CHPs, and HCs in multiple regions of Ethiopia.

## Methods

### Study design and setting

Ethiopia, located in horn of Africa, is administratively organized into 12 regional states, two city administrations, 92 zones, and more than 1,040 districts (*woredas*). The country’s healthcare system is structured into three levels of care: primary, secondary, and tertiary. At the primary level, a typical district serving about 60,000 to 100,000 people has a primary hospital, three to five health centers, and 20 health posts. In 2020, nationally 13014 (74%) of health posts were designated as BHPs, 1935 (11%) as CHPs, and 2638 (15%)as HEP Units [30].

The Improve Primary Health Care Service Delivery (IPHCSD) Project is a five-year initiative (2022-2027) implemented by Amref Health Africa and JSI Research & Training Institute Inc., in nine regional states of Ethiopia. The project aims to improve RMNCH health outcomes through implementation of HEPO Roadmap interventions, innovative primary healthcare service delivery models, evidence generation and prototype development for national scale-up. This facility-based, cross-sectional formative assessment was conducted between December 2024 and February 2025 to evaluate the implementation of HEPO-recommended RMNCH service package availability and readiness in restructured primary healthcare facilities. The study covered 139 ***woredas*** across 54 zones in eleven regional states and Dire Dawa City Administration, providing a broad assessment of HEPO implementation across diverse geographic and health system contexts in Ethiopia (Fig 1). This observational study was reported in accordance with the 22-item Strengthening the Reporting of Observational Studies in Epidemiology (STROBE) checklist [31].

**Figure 1.**
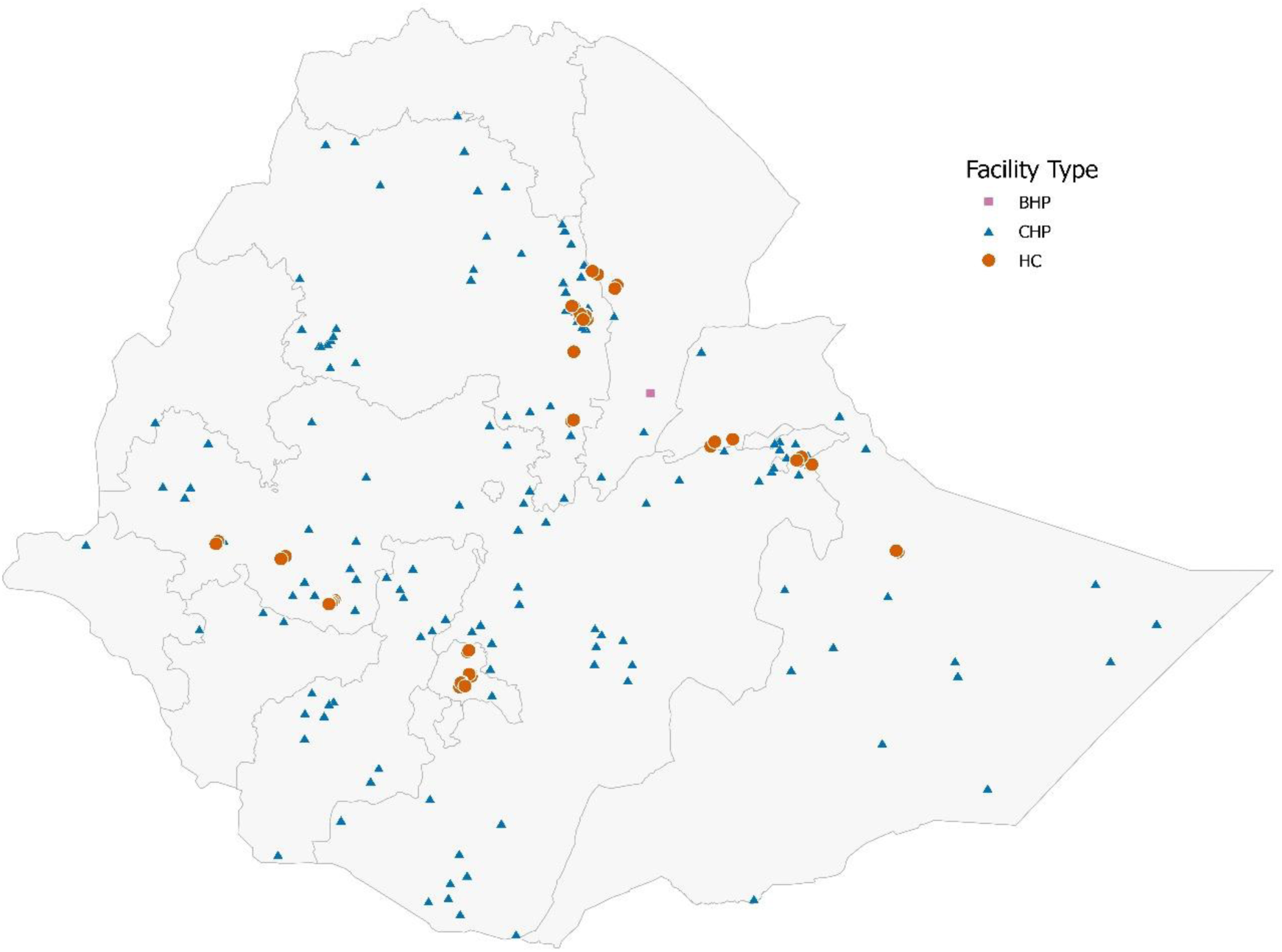
Geographic distribution of assessed BHPs, CHPs and HCs in Ethiopia, 2025.

### Study population and facility selection

The study targeted health facilities registered in the national routine health information system. All CHPs functional at the time of the survey and selected BHPs and HCs located in HEPO pilot and scale-up *woredas* were enrolled. The health facility was the unit of analysis. The final analytical sample comprised 305 primary health care facilities i.e., 107 BHPs, 142 CHPs and 56 HCs.

### Data collection instrument and quality assurance

Facility heads or designated officers provided interview data, and trained assessors directly observed tracer items at service-delivery points. A structured questionnaire was adapted from the WHO Service Availability and Readiness Assessment (SARA) and Ethiopian minimum requirement for each health facility [32]. The English-language instrument was programmed in KoboTool box with skip logic and legal-value constraints. It captured facility identification and context; governance and service organization; workforce; premises and amenities; equipment; infection-prevention supplies; medicines and commodities; and availability of family-planning, antenatal, childbirth, postnatal, under-five and malaria services.

Public-health professionals collected data after standardized training. Fifteen supervisors reviewed completeness and consistency daily and returned queries to field teams. The instrument was pretested and revised before fieldwork. Interviews with facility heads were complemented by direct observation of equipment, commodities, infection-prevention inputs, service rooms and recording tools. In addition, four years (2022-2025) of secondary health service data, including outpatient department (OPD) visits, and skilled birth attendance (SBA) services were extracted from the District Health Information System 2 (DHIS2) for analysis.

### Measures

#### General and service-specific readiness index

Health facilities readiness was assessed at two related but distinct levels. General facility readiness summarized five domains: human resources, premises/basic amenities, basic equipment, infection prevention/standard precautions, and medicines/commodities. General service availability was described separately and was not included in the five-domain general-readiness index. Service availability was analytically distinct from readiness. RMNCH service-availability scores were constructed for family planning, ANC, SBA, postnatal care and under-five care. Service availability measured whether a service or component was reported as offered, whereas readiness measured whether the inputs required to provide selected services were present. Domain scores were derived from the prespecified tracer items listed in the measurement framework. The tracer composition and denominator for each score are presented in Table 1.

**Table 1:** Description of domains and items used to construct health facility service availability and readiness assessment.

| Health Facility Readiness |  |
| --- | --- |
| Domains | Items |
| Human resource or staffing | 6 items: Midwives (all types) at least one present, Health Officer: at least one present, Nurse (all types), Pharmacist or pharmacy technician (all types) : at least one present, Laboratory technician or technologist (all types): at least one present, HEWs (level 3/level 4): at least one present. BHPs are required to have two HEWs |
| Domains | Items |
|  | and a Nurse. While both CHPs and HCs are expected to have all six health-worker categories. |
| Basic amenities | 23 items: External communication means, functioning facility-supported cellular phone, Functioning computer, Waiting area/room, MCH service room, labor room, PNC room, adult examination room, procedure room, drug/supply store or mini-dispensary, laboratory room, card/records room, staff office, adequate professional living rooms, guard room, janitor room, fenced compound, sex-separated/accessible latrine, access to electricity/power, functional backup generator/solar power, electricity available whenever facility was open, and water available for facility functions.<br>Basic Health Posts are required to have 19 of 23 items. Hence, no delivery, postnatal, procedure and laboratory rooms are required in BHPs. HCs are required to have 21 of 23 items. Hence, no staff living rooms are required in HCs. |
| Basic equipment | 8 items: Sterilizer (steam or dry heat), blood pressure apparatus, fetal stethoscope/ Pinard / doppler fetoscope, adult weighing scale, examination bed, measuring tape, cold box and refrigerator. All items are required in BHPs, CHPs and HCs. |
| Infection Prevention | 4 items: appropriate storage of sharps waste (sharps box/container), storage of infectious waste, availability of disinfectant and disposable latex gloves |
| Medicines | 5 items: amoxicillin dispersible tablet, oral rehydration salts, ferrous sulphate + folic acid, pentavalent vaccine, and Tetanus toxoid vaccine. All items are required in BHPs, CHPs and HCs |
| General Service Availability | 7 items: static health services, household services, community services, outreach services, school health services, workplace services, and health facility open 24/7. |
| RMNCH specific service availability |  |
| Family planning | 13 items: Offers FP services, Combined oral contraceptive pills, Progestin-only contraceptive pills, Combined injectable contraceptives, Progestin-only injectable contraceptives, Male condoms, Female condoms, Implants, Emergency contraceptive pills, Hormonal IUCD (Intrauterine Contraceptive Device), Non-hormonal IUCD, IUCD removal, and Implant removal.<br>Basic Health Posts are required to offer 9 of 13 items. Hence, no Hormonal IUCD Non-hormonal IUCD, IUCD removal, and Implant removal service required in BHPs. |
| Antenatal care | 12 items: Offers ANC services, Iron supplementation, Folic acid supplementation, Tetanus toxoid immunization, Blood-pressure measurement, Routine urine-protein testing, Human Immune Deficiency Virus (HIV) testing for pregnant women, Routine syphilis testing, Syphilis treatment, Diagnosis/treatment of other Sexually Transmitted Infections(STIs), Counseling to prevent Female genital mutilation (FGM), and Formal linkage with community health workers. All ANC service items are required to be offered in BHPs, CHPs and HCs. |
| Skilled Birth Attendance | 3 items: Offers Attendance or delivery care, offers Basic Emergency Obstetric and Newborn Care (BEmONC), delivery services available 24 hours. Basic Health Posts are not required to offer delivery service. |
| Postnatal care | 3 items: Routine outpatient maternal/newborn postnatal care, Maternal outpatient postnatal care, and formal linkage with community health workers, All Postnatal Care (PNC) service items are required to be offered in BHPs, CHPs and HCs. |
| Under-five care | 4 items: offers preventive/curative care for U5 children, Routine child growth monitoring, Diagnosis/treatment of child malnutrition and Outpatient Sever Acute Malnutrition (SAM) service with Ready-to-Use Therapeutic Food (RUTF). All U5 service items are required to be offered in BHPs, CHPs and HCs. |

#### Score construction

Tracer values were converted to a common 0–1 scale and domain scores were expressed from 0 to 100. Values of 1 indicated availability and values of 0 indicated absence. Within each domain, tracer values were summed, divided by the prespecified number of applicable tracers and multiplied by 100. The overall service-specific readiness index was the row mean of available ANC, SBA and under-five readiness scores. The general-readiness index was the equal-weighted mean of its five domains, while the RMNCH service-availability index was the equal-weighted mean of its five service domains. Structural absence and available-case scoring were examined when interpreting tier-specific comparisons.

### Statistical analysis

Statistical analyses were conducted using R statistical software version 4.6.0. Descriptive statistics were used to summarize the data. Categorical variables were summarized as frequencies and percentages. Continuous variables and scores were summarized using measures of central tendency and dispersion, including the mean, median, standard deviation (SD), and interquartile range (IQR). Data visualization was performed using bins, violin plots and box-and-whisker plots. Violin plots were used to illustrate the distribution of readiness scores across facility types, while box-and-whisker plots displayed the median and IQR; diamonds were used to indicate mean values. The normality assumption for continuous variables was assessed using the Kolmogorov-Smirnov test. Since the data significantly deviated from a normal distribution (p < 0.05), non-parametric statistical tests were applied. Differences in readiness scores among BHPs, CHPs, and HCs were compared using Kruskal–Wallis (H) tests for continuous variable scores [33] and Pearson chi-square or Fisher exact tests for categorical indicators [34], depending on cell sparsity. Kruskal–Wallis epsilon-squared was calculated as (H−k+1)/(N−k), where H is the test statistic, k is the number of facility groups and N is the number analyzed. Cramer’s V quantified the magnitude of association for categorical comparisons. The Benjamini–Hochberg procedure controlled the false discovery rate separately within prespecified families. The primary adjusted analysis used ordinary least-squares regression with the overall service-specific readiness index, expressed on a 0–100 scale, as the outcome. Prespecified covariates were facility type (CHP reference), urban–rural residence (urban reference), source/program region and log-transformed projected catchment population. Projected population was retained because completeness exceeded the prespecified 80% inclusion threshold. HC3 heteroskedasticity-robust standard errors were used. Regression coefficients are reported as adjusted percentage-point differences with 95% confidence intervals. All statistical tests were two-tailed, and p-values < 0.05 were considered statistically significant.

#### Validity and Reliability

Exploratory analyses examined the internal structure of the five-domain general-readiness measure. Cronbach’s alpha was 0.832 for human resources, 0.895 for basic amenities, 0.869 for basic equipment, 0.763 for medicines and commodities, and 0.630 for infection prevention. Alpha calculated across the five domain scores was 0.896. Four domains exceeded the conventional 0.70 threshold, whereas infection prevention fell below it [35]. Exploratory k-means clustering identified two facility-readiness profiles, with a silhouette coefficient of 0.469. Because SARA tracer inventories comprise distinct resources that together form a readiness index, these statistics describe patterns of co-availability and facility grouping rather than establish the index’s psychometric validity or reliability.

#### Ethical considerations

The protocol followed the principles of the Declaration of Helsinki and was approved by the Ethiopian Public Health Association Institutional Review Board (EPHA/OG/159/24). Administrative permission was obtained from regional, zonal and woreda health authorities. Facility respondents received information about the assessment, voluntary participation and withdrawal and provided written informed consent. Analytical data were maintained without patient-level information and handled confidentially.

## Results

### Facility characteristics

The study included 305 health facilities: 107 BHPs (35.1%), 142 CHPs (46.6%) and 56 HCs (18.4%). Oromia contributed 85 facilities (27.9%), Amhara 75 (24.6%), Somali 51 (16.7%) and Sidama 49 (16.1%). Most facilities were rural (269/305, 88.2%), including 99.1% of BHPs, 86.6% of CHPs and 71.4% of HCs (p<0.001) (Table 2). The distribution of health facilities also differed by contextual setting (p = 0.008). Overall, 16.7% of facilities were located in pastoralist areas and 83.3% in agrarian areas. CHPs had the lowest proportion of facilities situated in pastoralist settings (14.8%), compared with BHPs (29.9%) and HCs (32.1%) (Table 2).

**Table 2.** Characteristics of assessed BHPs, CHPs and HCs, Ethiopia, 2024–2025.

| Characteristics | BHP | Freq | CHP | Freq | HC | Freq | Total | Freq | p-value |
| --- | --- | --- | --- | --- | --- | --- | --- | --- | --- |
|  |  | (%) |  | (%) |  | (%) |  | (%) |  |
|  | (n=107) |  | (n=142) |  | (n=56) |  | (N=305) |  |  |
| Region |  |  |  |  |  |  |  |  |  |
| Afar Region | 12 (11.2) |  | 2 (1.4) |  | 6 (10.7) |  | 20 (6.6) |  | <0.001 |
| Amhara Region | 22 (20.6) |  | 42 (29.6) |  | 11 (19.6) |  | 75 (24.6) |  |  |
| Benishangul-Gumuz Region | 0 (0.0) |  | 1 (0.7) |  | 0 (0.0) |  | 1 (0.3) |  |  |
| Central Ethiopia Region | 0 (0.0) |  | 7 (4.9) |  | 0 (0.0) |  | 7 (2.3) |  |  |
| Dire Dawa City Administration | 0 (0.0) |  | 2 (1.4) |  | 0 (0.0) |  | 2 (0.7) |  |  |
| Gambella Region | 0 (0.0) |  | 2 (1.4) |  | 0 (0.0) |  | 2 (0.7) |  |  |
| Harari Region | 0 (0.0) |  | 2 (1.4) |  | 0 (0.0) |  | 2 (0.7) |  |  |
| Oromia Region | 22 (20.6) |  | 52 (36.6) |  | 11 (19.6) |  | 85 (27.9) |  |  |
| Sidama Region | 31 (29.0) |  | 2 (1.4) |  | 16 (28.6) |  | 49 (16.1) |  |  |
| Somali Region | 20 (18.7) |  | 19 (13.4) |  | 12 (21.4) |  | 51 (16.7) |  |  |
| South Ethiopia Region | 0 (0.0) |  | 9 (6.3) |  | 0 (0.0) |  | 9 (3.0) |  |  |
| South West Ethiopia Region | 0 (0.0) |  | 2 (1.4) |  | 0 (0.0) |  | 2 (0.7) |  |  |
| Location |  |  |  |  |  |  |  |  |  |
| Urban | 1 (0.9) |  | 19 (13.4) |  | 16 (28.6) |  | 36 (11.8) |  | <0.001 |
| Rural | 106 (99.1) |  | 123 (86.6) |  | 40 (71.4) |  | 269 (88.2) |  |  |
| Context |  |  |  |  |  |  |  |  |  |
| Pastoralist | 32(29.9) |  | 21(14.8) |  | 18(32.1) |  | 71 (23.3) |  | 0.008 |
| Agrarian | 75(70.1) |  | 121(85.2) |  | 38(67.9) |  | 234 (76.7) |  |  |

The overall median projected population served was 7,392 (IQR: 4,639-11,858). HCs served considerably larger populations, with a median catchment population of 25,000 (IQR: 18,389-29,973), compared with 6,531 (IQR: 4,639-9,331) for CHPs and 6,135 (IQR: 3,252-7,723) for BHPs (p < 0.001). Among CHPs, the median road distance to their supervising HC was 19.8 km (IQR: 9.0-25.3 km), ranging from 3 km to 83 km.

### General health facility readiness

Facility readiness increased progressively with the level of care. HCs achieved the highest mean general readiness score (86.1%), followed by CHPs (63.8%) and BHPs (33.2%). HCs performed particularly well in human resources (96.4%), medical equipment (87.7%), amenities (83.2%), medicines and commodities (83.6%), and infection-prevention capacity (79.5%). CHPs demonstrated relatively strong infection-prevention readiness (85.7%) and availability of medicines and commodities (82.3%) but had substantially lower human-resource readiness (17.6%). BHPs recorded the lowest scores across all readiness domains, especially human resources (7.5%) and basic amenities (15.9%). Figure 2 below demonstrates a pronounced readiness gradient by facility type, with the greatest gaps concentrated in human resources, amenities, and medical equipment at the primary-level facilities (**Supplementary Fig 1**).

**Figure 2.**
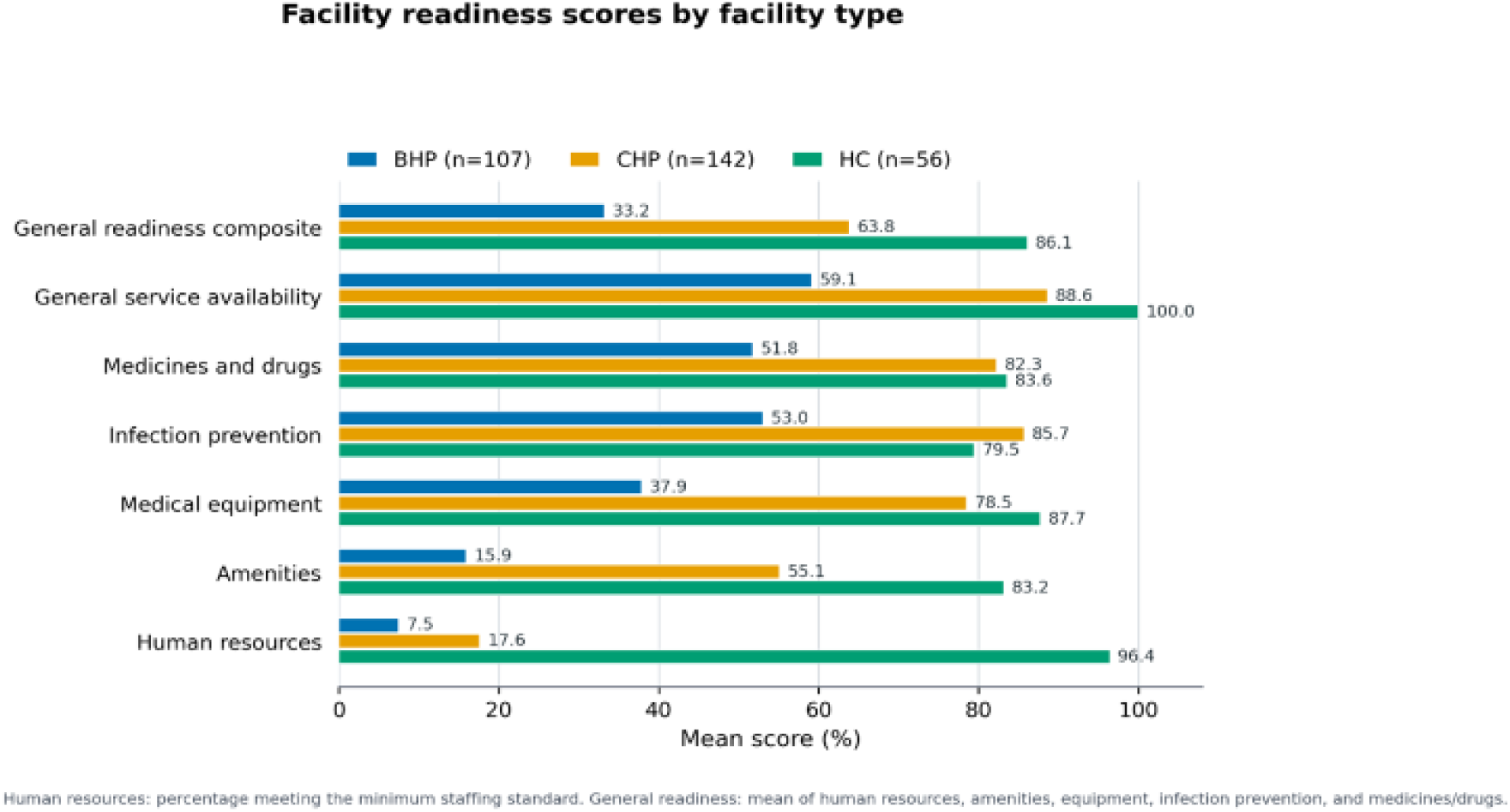
Bar charts depicting mean general health facility-readiness scores by domain and facility type Ethiopia, 2024–2025.

Table 3 presented substantial differences in general facility readiness across facility types. The median overall readiness index increased from 33.1% (IQR: 24.4–40.7) among BHPs to 62.9% (IQR: 54.1–71.4) among CHPs and 89.6% (IQR: 81.3–93.6) among HCs. This difference was statistically significant and had a large effect size (H(2)=204.53, p<0.001, ε²=0.673). Human-resource readiness was particularly limited at the lower facility levels: both BHPs and CHPs had median scores of 0.0% (IQR: 0.0–0.0), compared with 100.0% (IQR: 100.0–100.0) among HCs (H(2)=161.59, p<0.001, ε²=0.530). Median basic-amenities scores were 15.8% (IQR: 5.3–21.1), 56.5% (IQR: 39.1–69.6), and 85.7% (IQR: 76.2–90.5) for BHPs, CHPs, and HCs, respectively, representing the largest between-group variation (H(2)=216.57, p<0.001, ε²=0.711). Basic-equipment readiness followed a similar pattern, increasing from 37.5% (IQR: 25.0–50.0) among BHPs to 87.5% (IQR: 62.5–87.5) among CHPs and 93.8% (IQR: 87.5–100.0) among HCs (H(2)=158.20, p<0.001, ε²=0.517). CHPs had the highest median infection-prevention score at 100.0% (IQR: 75.0–100.0), compared with 87.5% (IQR: 50.0–100.0) among HCs and 50.0% (IQR: 25.0–75.0) among BHPs (H(2)=80.12, p<0.001, ε²=0.259). Medicines-readiness scores were 60.0% (IQR: 40.0–60.0) among BHPs and 80.0% among both CHPs and HCs (IQR: 80.0–100.0 for both), with significant variation across facility types (H(2)=128.16, p<0.001, ε²=0.418). Overall, readiness was consistently lowest among BHPs, while HCs generally achieved the highest scores across the assessed domains (**Supplementary file 1).**

**Table 3.** General facility-readiness scores by facility type.

| Domain | BHP Median<br>(IQR) | CHP Median<br>(IQR) | HC Median (IQR) | H(2) | p-value | Effect size<br>($\epsilon^2$) |
| --- | --- | --- | --- | --- | --- | --- |
| Human resources | 0.0 (0.0–0.0) | 0.0 (0.0–0.0) | 100.0 (100.0–100.0) | 161.59 | <0.001 | 0.530 |
| Basic amenities | 15.8 (5.3–21.1) | 56.5 (39.1–69.6) | 85.7 (76.2–90.5) | 216.57 | <0.001 | 0.711 |
| Basic equipment | 37.5 (25.0–50.0) | 87.5 (62.5–87.5) | 93.8 (87.5–100.0) | 158.20 | <0.001 | 0.517 |
| Infection prevention | 50.0 (25.0–75.0) | 100.0 (75.0–100.0) | 87.5 (50.0–100.0) | 80.12 | <0.001 | 0.259 |
| Medicines | 60.0 (40.0–60.0) | 80.0 (80.0–100.0) | 80.0 (80.0–100.0) | 128.16 | <0.001 | 0.418 |
| **Overall index** | **33.1 (24.4–40.7)** | **62.9 (54.1–71.4)** | **89.6 (81.3–93.6)** | **204.53** | **<0.001** | **0.673** |

### Service-specific readiness RMNCH service availability

Service-specific readiness differed significantly across facility types (Figure 3). Median overall readiness increased from 67.9% (IQR: 56.4–68.5) among BHPs to 77.8% (IQR: 60.0–86.8) among CHPs and 91.1% (IQR: 85.4–94.4) among HCs (H(2)=49.15, p<0.001, ε²=0.236). The largest between-facility difference was observed for ANC readiness: median scores were 50.0% (IQR: 40.0–60.0), 80.0% (IQR: 70.0–90.0) and 95.0% (IQR: 90.0–100.0), respectively (H(2)=179.71, p<0.001, ε²=0.588). Childbirth-readiness medians were 45.5% (IQR: 27.3–63.6) among the five BHPs with childbirth data, 58.3% (IQR: 0.0–75.0) among CHPs and 83.3% (IQR: 75.0–91.7) among HCs (H(2)=38.63, p<0.001, ε²=0.183). Although median under-five readiness was 100.0% in all facility types, BHPs had a wider distribution (IQR: 75.0–100.0) than CHPs and HCs (IQR:100.0–100.0), producing a statistically significant but comparatively small difference (H(2)=27.56, p<0.001, ε²=0.085) (**Supplementary file 2**).

**Figure 3.**
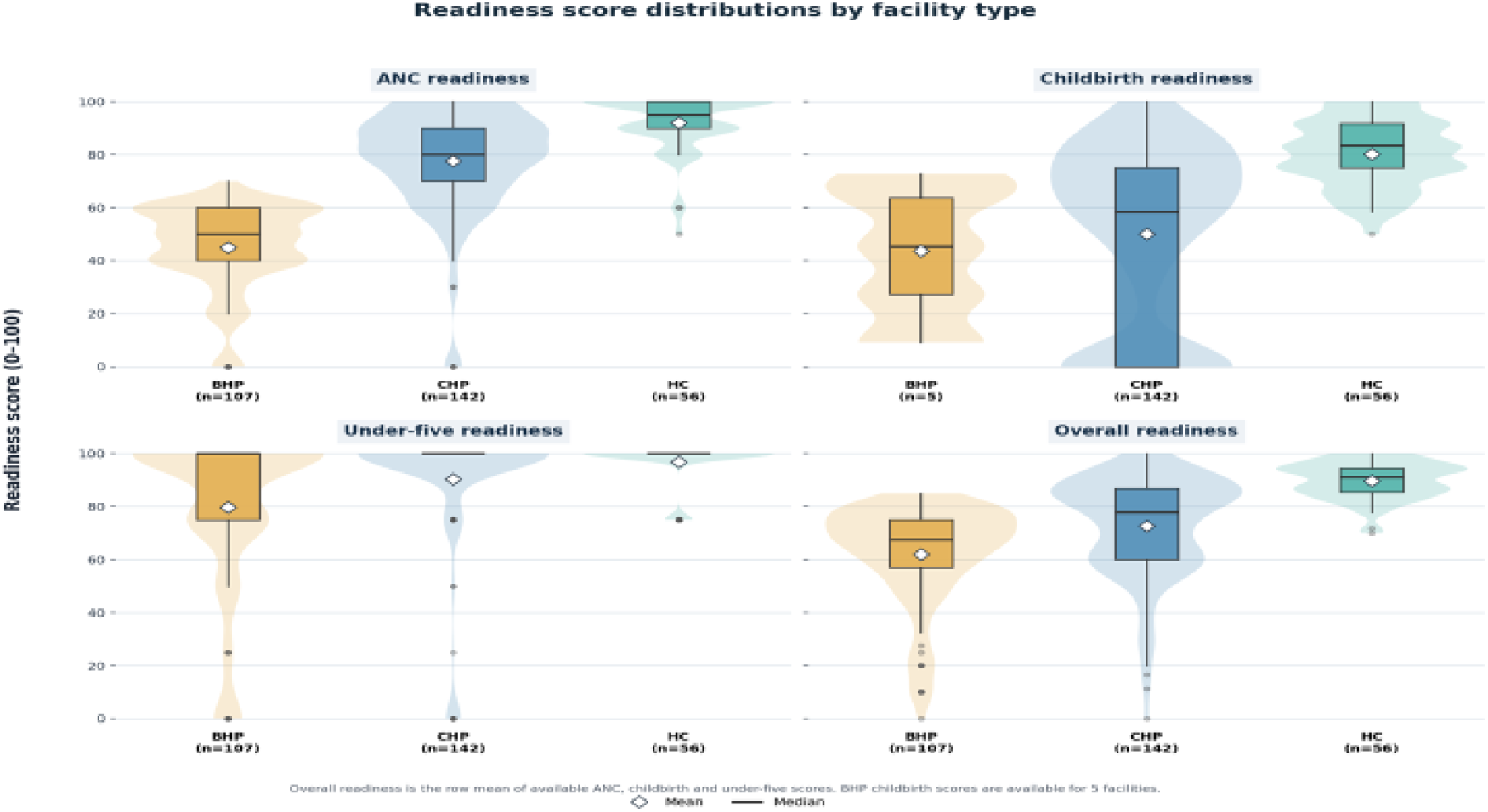
Distribution of ANC, childbirth, under-five and overall service-specific readiness scores by facility type, Ethiopia, 2024–2025

RMNCH service availability varied significantly across facility types. The median overall service-availability score increased from 71.8% (IQR: 67.2–76.6) among basic health posts (BHPs) to 79.0% (IQR: 67.8–86.0) among comprehensive health posts (CHPs) and 88.0% (IQR: 81.8–92.3) among health centers (HCs) (H(2)=31.08, p<0.001, ε²=0.145). The most pronounced difference was observed for antenatal care, with median scores of 50.0%, 80.0%, and 95.0% among BHPs, CHPs, and HCs, respectively (H(2)=179.71, p<0.001, ε²=0.588). Skilled-birth-attendance scores also increased across facility levels, from 45.5% among BHPs to 58.3% among CHPs and 83.3% among HCs (H(2)=38.63, p<0.001, ε²=0.183). Family Planning scores were comparatively similar 66.7% at BHPs and 69.2% at both CHPs and HCs and the observed difference had a small effect size (H(2)=6.57, p=0.037, ε²=0.015). Although the median scores for postnatal and under-five care were 100.0% across all facility types, the wider distributions among BHPs produced statistically significant differences (p<0.001), with small effect sizes of 0.048 and 0.085, respectively. Overall, HCs demonstrated the highest RMNCH service availability, with the greatest facility-level disparities occurring in antenatal care and skilled birth attendance(Table 4).

**Table 4.** RMNCH service-availability scores by facility type.

| Domain | BHP Median (IQR) | CHP Median [IQR] | HC Median [IQR] | H(2) | p-value | Effect size ($\epsilon^2$) |
| --- | --- | --- | --- | --- | --- | --- |
| Family planning | 66.7 (55.6–77.8) | 69.2 (53.8–76.9) | 69.2 (61.5–76.9) | 6.57 | 0.037 | 0.015 |
| Antenatal care | 50.0 (40.0–60.0) | 80.0 (70.0–90.0) | 95.0 (90.0–100.0) | 179.71 | <0.001 | 0.588 |
| Skilled Birth Attendance | 45.5 (27.3–63.6) | 58.3 (0.0–75.0) | 83.3 (75.0–91.7) | 38.63 | <0.001 | 0.183 |
| Postnatal care | 100.0 (50.0–100.0) | 100.0 (100.0–100.0) | 100.0 (100.0–100.0) | 16.51 | <0.001 | 0.048 |
| Under-five care | 100.0 (75.0–100.0) | 100.0 (100.0–100.0) | 100.0 (100.0–100.0) | 27.56 | <0.001 | 0.085 |
| Overall index | **71.8 (67.2–76.6)** | **79.0 (67.8–86.0)** | **88.0 (81.8–92.3)** | **31.08** | **<0.001** | **0.145** |

### Health service volumes offered by facility type

Over the four-year study period (2022-2025), outpatient service utilization increased substantially following the upgrading of BHPs to CHPs. On average, outpatient service volume increased by approximately 180 visits per facility per year after the transition. The findings indicate that outpatient visits in CHPs nearly tripled compared with pre-upgrading levels and, among facilities providing services for more than 12 months, approached the service volumes observed in health centers (Fig 4).

**Figure 4.**
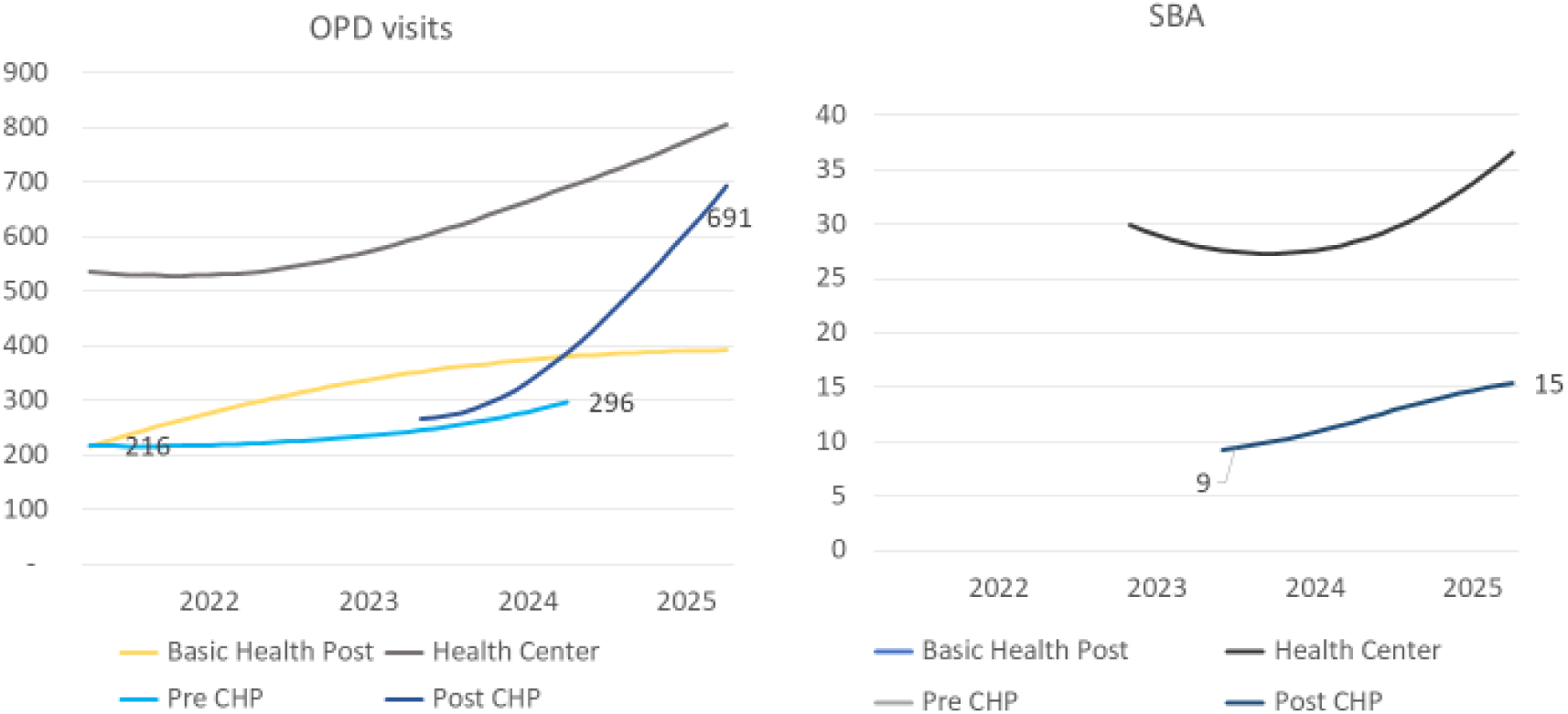
Line charts depicting monthly service volume per facility type, 2022-2025

Similarly, utilization of skilled delivery services increased markedly after the introduction of CHPs. The average number of skilled deliveries rose from 9 deliveries per CHP per month to 15 deliveries per month, representing an increase of approximately 67%. This finding suggests a substantial expansion in access to and uptake of facility-based maternity services following facility upgrading (Fig 4). Overall, the line charts demonstrate that upgrading BHPs to CHPs was associated with substantial increases in the utilization of outpatient, and maternity services, with service volumes progressively approaching those observed in established health centers.

### Adjusted associations with overall service-specific readiness

In the adjusted analysis, facility type and projected catchment population were significantly associated with overall service-specific readiness (Table 5). Using CHPs as the reference group, BHPs scored 9.9 percentage points lower (adjusted difference: −9.9; robust SE: 2.4; 95% CI: −14.7 to −5.1; p<0.001), whereas HCs scored 8.7 points higher (adjusted difference: 8.7; robust SE: 3.6; 95% CI: 1.7–15.8; p=0.015). Rural location was not independently associated with readiness compared with urban location (adjusted difference: 2.4; robust SE: 3.3; 95% CI: −4.1 to 8.8; p=0.476). Each one-unit increase in log-transformed projected population was associated with a 6.6-point increase in overall service-specific readiness (robust SE: 2.3; 95% CI: 2.1–11.1; p=0.004).

**Table 5.**
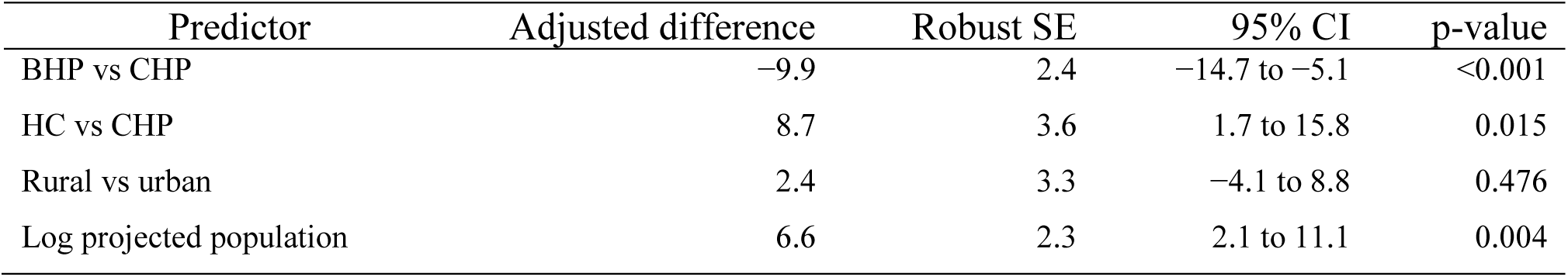
Adjusted associations with overall service-specific readiness.

| Predictor | Adjusted difference | Robust SE | 95% CI | p-value |
| --- | --- | --- | --- | --- |
| BHP vs CHP | -9.9 | 2.4 | -14.7 to -5.1 | <0.001 |
| HC vs CHP | 8.7 | 3.6 | 1.7 to 15.8 | 0.015 |
| Rural vs urban | 2.4 | 3.3 | -4.1 to 8.8 | 0.476 |
| Log projected population | 6.6 | 2.3 | 2.1 to 11.1 | 0.004 |

## Discussion

This assessment of RMNCH service availability and health facility readiness across 11 regional states of Ethiopia demonstrated substantial variation among BHPs, CHPs, and HCs. Consistent with the Health Extension Program Optimization (HEPO) roadmap, most CHPs were located in rural and underserved areas, often more than 90 minutes’ walking distance from their supervising health centers, highlighting their strategic role in expanding access to primary healthcare services and extending the reach of the primary healthcare system to remote populations [18,36]. The findings indicate that CHPs occupy an intermediate position between BHPs and HCs in terms of service capacity and utilization, contributing to improved access to outpatient care, skilled birth attendance, and a range of RMNCH services for populations with limited access to facility-based care [18,37]. Despite these gains, significant gaps remain in the deployment of the required health workforce, availability of essential infrastructure, and overall compliance with national minimum standards for both BHPs and CHPs [36,38]. These limitations may constrain the ability of lower-level facilities to deliver the full package of essential RMNCH services and achieve the intended goals of the HEPO reform and universal health coverage agenda in Ethiopia [18, 38].

Only one in ten BHPs met the HEO recommendations on staffing each facility with two HEWs (HEW and one nurse, indicating a major frontline health workforce deficit. HEPO was designed to strengthen primary healthcare through an expanded and more skilled workforce capable of delivering comprehensive promotive, preventive, and curative services at community level [18]. The low level of compliance observed in this study suggests that implementation of the workforce reforms has not yet achieved the intended scale. Similar challenges related to recruitment, deployment, retention, and equitable distribution of health workers have been reported across Ethiopia’s primary healthcare system, limiting service delivery capacity and health system performance. Inadequate staffing at BHPs may compromise the availability, quality, and continuity of RMNCH services, particularly in rural and underserved areas where communities rely heavily on primary healthcare facilities. Furthermore, although CHPs are expected to function through multidisciplinary teams [18], fewer than one-fifth of CHPs had the full complement of required personnel. This finding suggests that the transition from conventional health posts to comprehensive community-based primary care units remains incomplete. The absence of multidisciplinary teams may hinder the delivery of integrated RMNCH, nutrition, communicable disease, and non-communicable disease services. Consistent with WHO recommendations, an adequately staffed and multidisciplinary primary healthcare workforce is essential for improving service readiness, accessibility, continuity of care, and progress toward UHC [39]. These findings underscore the need for strengthened workforce planning, recruitment, deployment, and retention strategies to achieve the objectives of the HEPO roadmap and improve primary healthcare service readiness in Ethiopia [18, 36, 39].

The principal service-specific gaps were in ANC and childbirth. Although childbirth readiness increased with facility level, the CHP median of 58.3% indicates that assigning a facility a childbirth mandate did not consistently ensure the full set of inputs needed for safe care. This pattern is consistent with Ethiopian studies documenting gaps in reliable electricity and water, essential medicines, newborn-resuscitation equipment, staff training and emergency transport at primary-care facilities [20,28,29]. The finding was especially important because Ethiopia’s national policy framework seeks universal performance of basic emergency obstetric and newborn care signal functions, while the Essential Health Services Package places ANC, SBA, immediate newborn care and PNC within an integrated continuum of care [36,37,40,41].

ANC readiness was substantially lower in BHPs than in CHPs and HCs. This result is consistent with national evidence showing shortfalls in trained staff and guidelines, diagnostics, medicines and infection-prevention inputs [42]. National standards require ANC to include more than registration of a visit: blood-pressure assessment, screening and testing, preventive medicines, counselling and timely referral must be reliably available. Consequently, routine monitoring should not classify an ANC platform as functional solely because the service is reported as offered; it should verify the tracer package and the care processes delivered [43].

Family-planning service availability differed little across facility types and had a very small effect size, but median scores around 67%–69% indicate an incomplete method mix. This finding should be interpreted against the national commitment to informed choice and equitable access to short- and long-acting FP methods in the Essential Health Services Package [36,40]. HEPO implementation should therefore protect uninterrupted contraceptive supply, provider competency, counselling quality and both insertion and removal capacity for long-acting reversible methods. Reporting the availability of any family-planning service alone can mask limitations that constrain women’s ability to choose and continue a preferred method [44].

Postnatal- and under-five-service availability reached median ceiling values, but ceiling effects do not demonstrate comprehensive quality. Ethiopian guidance calls for mothers and newborns to remain under facility care for at least 24 hours after an uncomplicated birth and requires early postnatal assessment, counselling, identification of danger signs and referral [39]. Similarly, under-five services depend on functioning assessment tools, essential medicines, nutrition services and referral systems. The wider score distributions among BHPs and persistent general-readiness deficits in amenities, infection prevention and medicines indicate that nominal availability must be interpreted alongside the inputs and clinical processes required for safe care [25,26,45].

The adjusted comparisons have direct implications for the HEPO roadmap. CHPs occupied an intermediate position and performed better than BHPs, but they remained below HCs after adjustment. This supports the HEPO principle of differentiating platforms by geography and service mandate while showing that reclassification alone is insufficient [17]. Upgrading a BHP to a CHP should be treated as an operational package: the required workforce mix, staff accommodation, water, power, rooms, equipment, diagnostics, medicines, infection-prevention systems, supervision and referral arrangements should be funded and deployed together. Facility designation should be contingent on verified readiness to deliver the assigned package safely.

### Policy Implication of RMNCH service availability and facility readiness assessment

The main purpose of this study was to determine RMNCH service availability and health facility readiness after endorsing HEPO roadmap with restructured primary healthcare facilities with practices, premises, professionals and products. The Ministry of Health and Institute of Ethiopia Standards should prioritize three linked actions. First, childbirth readiness should be managed as a complete safety bundle that includes round-the-clock skilled coverage, reliable water and power, uterotonics and magnesium sulphate, maternal monitoring, newborn resuscitation, infection prevention, communication and emergency transport. Second, regional health bureaus and woredas should use a tier-specific certification checklist before commissioning or renewing childbirth services, with time-bound corrective plans for unmet standards. Third, procurement, workforce deployment and capital investment should use the same facility registry and readiness dashboard so that new mandates are not introduced without the resources required to perform them.

These recommendations align with Ethiopia’s Health Sector Transformation Plan II and National Healthcare Quality and Safety Strategy, which emphasize equitable, people-centred care, clinical safety, accountability and continuous improvement [37,38, 46, 47]. Routine monitoring should therefore report service availability and readiness separately and add measures of effective coverage, including adherence to clinical standards, referral completion, respectful care, patient experience and maternal and newborn outcomes. Readiness results should be reviewed through PHCU and woreda performance systems, with supportive supervision and financing directed to the specific standards that facilities fail to meet rather than to undifferentiated infrastructure expansion.

### Strengths and limitations

Strengths of this assessment include direct comparison of three restructured primary healthcare facilities, inclusion of multiple regions and service contexts, direct observation of tracer inputs, explicit score definitions, robust inference and separation of general readiness, RMNCH service availability and service-specific readiness. Distinguishing these constructs reduced the risk of treating nominal service provision as evidence that all required inputs were present.

Several limitations warrant careful interpretation. First, this formative assessment was conducted during the first two years of HEPO roadmap implementation; consequently, SARA domain scores may change as implementation matures and targeted interventions are introduced. Second, facilities were selected from registered CHPs and project-participating woredas. Because national selection probabilities and sampling weights were unavailable, the findings should not be interpreted as national prevalence estimates. Third, the cross-sectional design precludes causal attribution of observed differences to facility restructuring. Residual confounding by financing, remoteness, management capacity, supervision, supply-chain performance, and service volume may remain. Moreover, adjustment for catchment population may partially control for factors closely related to facility designation and resource allocation; the adjusted estimates should therefore be interpreted as conditional associations rather than total effects of facility type. Fourth, service mandates differ across facility tiers, and lower readiness scores may partly reflect an intentionally narrower service scope rather than deficiencies in expected functions. Childbirth-readiness data were available for only five BHPs, resulting in substantial imprecision and limiting direct comparisons with CHPs and health centers. The overall service-specific readiness score was calculated from available domain scores and should not be interpreted as an all-facility composite in which every structurally unavailable service was assigned a score of zero. Finally, a single assessment visit cannot capture the duration of stock-outs, provider competence, care processes, respectful care, patient experience, referral completion, service continuity, or clinical outcomes.

### Conclusions

Among the assessed Ethiopian facilities, CHPs expanded the service package beyond BHPs but did not consistently attain HC readiness. Major gaps remained in staffing, amenities, equipment and childbirth capacity. HEPO scale-up should link each facility designation to enforceable tier-specific minimum standards, synchronized workforce and commodity deployment, verified childbirth-safety bundles, competency-based supervision and reliable emergency referral. National and subnational monitoring should distinguish availability from readiness and assess whether restructuring improves effective coverage, equity and maternal and child health outcomes.

## Declarations

## Abbreviations

ANC: Antenatal care
BEmONC: Basic Emergency Obstetric and Newborn Care
BHP: Basic Health Post
CHP: Comprehensive Health Post
CI: Confidence Interval
HC: Health Center
HEP: Health Extension Program
HEPO: Health Extension Program Optimization Roadmap
HEW: Health Extension Worker
IPHCSD: Improve Primary Health Care Service Delivery
IQR: Interquartile Range
IUCD: Intrauterine contraceptive Device
PHC: Primary Healthcare
PHCU: Primary Healthcare Unit
PNC: Postnatal Care
RMNCH: Reproductive, Maternal, Newborn, and Child Health
SARA: Service Availability and Readiness Assessment
SBA: Skilled Birth Attendance
SD: Standard Deviation
SDGs: Sustainable Development Goals
UHC: Universal Health Coverage
WHO: World Health Organization

## Ethical consideration

The research protocol of this retrospective cross-sectional facility-based study design was reviewed by the Ethiopian Public Health Association (EPHA) research ethical review board (Ref no. EPHA/OG/159/24). The study was conducted by the Declaration of Helsinki. Informed verbal and written consent were obtained from each study participant voluntarily to be included in the study. Permission was sought to use the anonymous project data from Amref Health Africa. The collected data was kept confidential anonymously through the de-identification of names and other personal identifiers from the record/sheet.

## Consent for publication

Not applicable

## Availability of data and materials

All related data are presented fully within the paper, and available upon reasonable request to the lead author and the corresponding author.

## Use of AI tools

During the preparation of this manuscript, the authors used ChatGPT-6 Sol to assist with chart development and English language editing. The authors reviewed and revised all AI-assisted content and take full responsibility for the accuracy and integrity of the published work.

## Authors’ contributions

MDA, GM, AS, GTT, MB, MT, SMT, DTA, SM, YK, AK, RD, WN, AG, YW, FS, BTT, TA TAB, IAO, AT, LT and BH have been involved during the inception of the study design the formative assessment of HEPO in general and RMNCH service availability and health facilities readiness after restructuring national exercise specifically. MDA, AS, RD, YK, WN, AG, AT, IAO, DTA, SM and FS are involved in the data acquisition and quality assurance measures. MDA*, GM, AS, GTT, and DTA carried out the data analysis and interpretation of the findings. MDA*, GM, AS, MT, GTT, and GTT developed the manuscript, and MT, MB, AS, GM, GTT, RD, YW, AG, SMT, and TAB reviewed the manuscript for intellectual content. AT, HW, MDM, LT, FD, KLH, LT, DE, MK, TRM and BH gave high-level critical comments and oversaw project implementation. All authors have reviewed the manuscript critically for important intellectual content and approved the final manuscript for submission.

## Acknowledgments

We gratefully acknowledge the Ministry of Health, The Gates Foundation and its Institute for Disease Modeling (IDM) team for their financial and technical support. We also thank Amref Health Africa for its overall administrative support and the study participants, data collectors, and supervisors for generously contributing their time, knowledge, and expertise to this study.

## Supporting information

**S1 Table 1.**
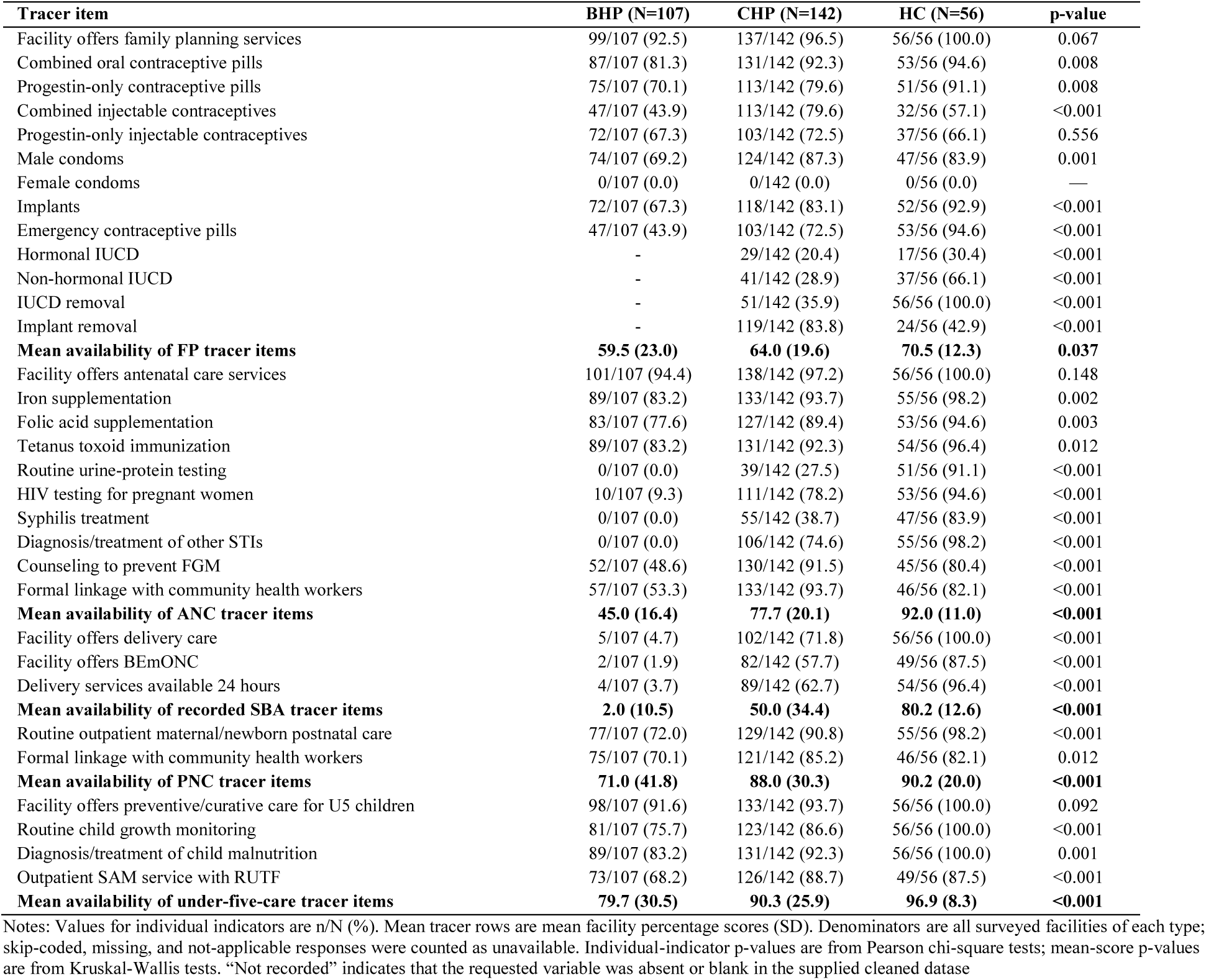
RMNHC service availability by facility type.

| Tracer item | BHP (N=107) | CHP (N=142) | HC (N=56) | p-value |
| --- | --- | --- | --- | --- |
| Facility offers family planning services | 99/107 (92.5) | 137/142 (96.5) | 56/56 (100.0) | 0.067 |
| Combined oral contraceptive pills | 87/107 (81.3) | 131/142 (92.3) | 53/56 (94.6) | 0.008 |
| Progestin-only contraceptive pills | 75/107 (70.1) | 113/142 (79.6) | 51/56 (91.1) | 0.008 |
| Combined injectable contraceptives | 47/107 (43.9) | 113/142 (79.6) | 32/56 (57.1) | <0.001 |
| Progestin-only injectable contraceptives | 72/107 (67.3) | 103/142 (72.5) | 37/56 (66.1) | 0.556 |
| Male condoms | 74/107 (69.2) | 124/142 (87.3) | 47/56 (83.9) | 0.001 |
| Female condoms | 0/107 (0.0) | 0/142 (0.0) | 0/56 (0.0) | — |
| Implants | 72/107 (67.3) | 118/142 (83.1) | 52/56 (92.9) | <0.001 |
| Emergency contraceptive pills | 47/107 (43.9) | 103/142 (72.5) | 53/56 (94.6) | <0.001 |
| Hormonal IUCD | - | 29/142 (20.4) | 17/56 (30.4) | <0.001 |
| Non-hormonal IUCD | - | 41/142 (28.9) | 37/56 (66.1) | <0.001 |
| IUCD removal | - | 51/142 (35.9) | 56/56 (100.0) | <0.001 |
| Implant removal | - | 119/142 (83.8) | 24/56 (42.9) | <0.001 |
| **Mean availability of FP tracer items** | **59.5 (23.0)** | **64.0 (19.6)** | **70.5 (12.3)** | **0.037** |
| Facility offers antenatal care services | 101/107 (94.4) | 138/142 (97.2) | 56/56 (100.0) | 0.148 |
| Iron supplementation | 89/107 (83.2) | 133/142 (93.7) | 55/56 (98.2) | 0.002 |
| Folic acid supplementation | 83/107 (77.6) | 127/142 (89.4) | 53/56 (94.6) | 0.003 |
| Tetanus toxoid immunization | 89/107 (83.2) | 131/142 (92.3) | 54/56 (96.4) | 0.012 |
| Routine urine-protein testing | 0/107 (0.0) | 39/142 (27.5) | 51/56 (91.1) | <0.001 |
| HIV testing for pregnant women | 10/107 (9.3) | 111/142 (78.2) | 53/56 (94.6) | <0.001 |
| Syphilis treatment | 0/107 (0.0) | 55/142 (38.7) | 47/56 (83.9) | <0.001 |
| Diagnosis/treatment of other STIs | 0/107 (0.0) | 106/142 (74.6) | 55/56 (98.2) | <0.001 |
| Counseling to prevent FGM | 52/107 (48.6) | 130/142 (91.5) | 45/56 (80.4) | <0.001 |
| Formal linkage with community health workers | 57/107 (53.3) | 133/142 (93.7) | 46/56 (82.1) | <0.001 |
| **Mean availability of ANC tracer items** | **45.0 (16.4)** | **77.7 (20.1)** | **92.0 (11.0)** | **<0.001** |
| Facility offers delivery care | 5/107 (4.7) | 102/142 (71.8) | 56/56 (100.0) | <0.001 |
| Facility offers BEmONC | 2/107 (1.9) | 82/142 (57.7) | 49/56 (87.5) | <0.001 |
| Delivery services available 24 hours | 4/107 (3.7) | 89/142 (62.7) | 54/56 (96.4) | <0.001 |
| **Mean availability of recorded SBA tracer items** | **2.0 (10.5)** | **50.0 (34.4)** | **80.2 (12.6)** | **<0.001** |
| Routine outpatient maternal/newborn postnatal care | 77/107 (72.0) | 129/142 (90.8) | 55/56 (98.2) | <0.001 |
| Formal linkage with community health workers | 75/107 (70.1) | 121/142 (85.2) | 46/56 (82.1) | 0.012 |
| **Mean availability of PNC tracer items** | **71.0 (41.8)** | **88.0 (30.3)** | **90.2 (20.0)** | **<0.001** |
| Facility offers preventive/curative care for U5 children | 98/107 (91.6) | 133/142 (93.7) | 56/56 (100.0) | 0.092 |
| Routine child growth monitoring | 81/107 (75.7) | 123/142 (86.6) | 56/56 (100.0) | <0.001 |
| Diagnosis/treatment of child malnutrition | 89/107 (83.2) | 131/142 (92.3) | 56/56 (100.0) | 0.001 |
| Outpatient SAM service with RUTF | 73/107 (68.2) | 126/142 (88.7) | 49/56 (87.5) | <0.001 |
| **Mean availability of under-five-care tracer items** | **79.7 (30.5)** | **90.3 (25.9)** | **96.9 (8.3)** | **<0.001** |

Notes: Values for individual indicators are n/N (%). Mean tracer rows are mean facility percentage scores (SD). Denominators are all surveyed facilities of each type; skip-coded, missing, and not-applicable responses were counted as unavailable. Individual-indicator p-values are from Pearson chi-square tests; mean-score p-values are from Kruskal-Wallis tests. "Not recorded" indicates that the requested variable was absent or blank in the supplied cleaned dataset

**S1 Table 2.** Facility Readiness by facility type.

| Tracer item | BHP (N=107) | CHP (N=142) | HC (N=56) | p-value |
| --- | --- | --- | --- | --- |
| Midwives (all types): at least one present | -) | 107/142 (75.4) | 55/56 (98.2) | <0.001 |
| Health officers: at least one present | - | 66/142 (46.5) | 55/56 (98.2) | <0.001 |
| Nurses (all types): at least one present | 20/107 (18.7) | 132/142 (93.0) | 55/56 (98.2) | <0.001 |
| Pharmacy staff: at least one present | - | 68/142 (47.9) | 54/56 (96.4) | <0.001 |
| Laboratory staff: at least one present | - | 42/142 (29.6) | 55/56 (98.2) | <0.001 |
| Health extension workers: at least one present | 100/107 (93.5) | 136/142 (95.8) | 52/56 (92.9) | 0.624 |
| **Mean availability of HR -six recorded staff-cadre tracers** | **18.7 (6.4)** | **64.7 (23.1)** | **97.0 (6.4)** | **<0.001** |
| Functioning facility-supported cellular phone | 24/107 (22.4) | 51/142 (35.9) | 20/56 (35.7) | 0.054 |
| Functioning computer | 2/107 (1.9) | 15/142 (10.6) | 48/56 (85.7) | <0.001 |
| Waiting area/room | 40/107 (37.4) | 101/142 (71.1) | 51/56 (91.1) | <0.001 |
| MCH service room | 22/107 (20.6) | 131/142 (92.3) | 56/56 (100.0) | <0.001 |
| Labor room | - | 90/142 (63.4) | 56/56 (100.0) | <0.001 |
| PNC room | - | 91/142 (64.1) | 56/56 (100.0) | <0.001 |
| Adult examination room | 22/107 (20.6) | 129/142 (90.8) | 55/56 (98.2) | <0.001 |
| Procedure room | 0/107 (0.0) | 95/142 (66.9) | 38/56 (67.9) | <0.001 |
| Drug/supply store or mini-dispensary | 45/107 (42.1) | 124/142 (87.3) | 56/56 (100.0) | <0.001 |
| Laboratory room | - | 60/142 (42.3) | 54/56 (96.4) | <0.001 |
| Card/records room | - | 81/142 (57.0) | 52/56 (92.9) | <0.001 |
| Staff office | 5/107 (4.7) | 66/142 (46.5) | 42/56 (75.0) | <0.001 |
| Professional living rooms | 14/107 (13.1) | 77/142 (54.2) | - | <0.001 |
| Adequate professional living rooms | 0/107 (0.0) | 39/142 (27.5) | - | <0.001 |
| Guard room | 20/107 (18.7) | 94/142 (66.2) | 39/56 (69.6) | <0.001 |
| Janitor room | 0/107 (0.0) | 33/142 (23.2) | 56/56 (100.0) | <0.001 |
| Fenced compound | 5/107 (4.7) | 92/142 (64.8) | 56/56 (100.0) | <0.001 |
| Sex-separated/accessible latrine | 5/107 (4.7) | 142/142 (100.0) | 56/56 (100.0) | <0.001 |
| Access to electricity/power | 12/107 (11.2) | 61/142 (43.0) | 42/56 (75.0) | <0.001 |
| Functional backup generator/solar power | 42/107 (39.3) | 34/142 (23.9) | 26/56 (46.4) | 0.003 |
| Electricity available whenever facility was open | 25/107 (23.4) | 46/142 (32.4) | 21/56 (37.5) | 0.128 |
| Water available for facility functions | 41/107 (38.3) | 65/142 (45.8) | 42/56 (75.0) | <0.001 |
| **Mean availability of basic-amenity tracer items** | **15.9 (11.4)** | **55.1 (20.2)** | **83.2 (9.2)** | **<0.001** |
| Sterilizer (steam or dry heat) | 5/107 (4.7) | 64/142 (45.1) | 42/56 (75.0) | <0.001 |
| Blood-pressure apparatus | 43/107 (40.2) | 127/142 (89.4) | 48/56 (85.7) | <0.001 |
| Fetal stethoscope/ Pinnard/Doppler | 48/107 (44.9) | 126/142 (88.7) | 46/56 (82.1) | <0.001 |
| Adult weighing scale | 47/107 (43.9) | 108/142 (76.1) | 50/56 (89.3) | <0.001 |
| Examination bed | 68/107 (63.6) | 118/142 (83.1) | 53/56 (94.6) | <0.001 |
| Tape measure | 51/107 (47.7) | 97/142 (68.3) | 47/56 (83.9) | <0.001 |
| Delivery bed with stirrups | 3/107 (2.8) | 97/142 (68.3) | 51/56 (91.1) | <0.001 |
| Examination light | 1/107 (0.9) | 36/142 (25.4) | 38/56 (67.9) | <0.001 |
| Cord clamp | 3/107 (2.8) | 90/142 (63.4) | 52/56 (92.9) | <0.001 |
| Episiotomy scissors | 2/107 (1.9) | 83/142 (58.5) | 53/56 (94.6) | <0.001 |
| Outlet forceps | 1/107 (0.9) | 66/142 (46.5) | 24/56 (42.9) | <0.001 |
| Pulse oximeter | 0/107 (0.0) | 22/142 (15.5) | 19/56 (33.9) | <0.001 |
| Cold box with ice packs | 57/107 (53.3) | 124/142 (87.3) | 51/56 (91.1) | <0.001 |
| **Mean availability of basic-equipment tracer items** | **23.7 (14.5)** | **62.7 (21.7)** | **78.8 (15.2)** | **<0.001** |
| Appropriate sharps-waste storage | 69/107 (64.5) | 125/142 (88.0) | 37/56 (66.1) | <0.001 |
| Appropriate infectious-waste storage | 27/107 (25.2) | 107/142 (75.4) | 37/56 (66.1) | <0.001 |
| Disinfectants available | 52/107 (48.6) | 123/142 (86.6) | 52/56 (92.9) | <0.001 |
| Disposable latex gloves | 79/107 (73.8) | 132/142 (93.0) | 52/56 (92.9) | <0.001 |
| **Mean availability of infection-prevention tracer items** | **53.0 (29.3)** | **85.7 (23.3)** | **79.5 (23.9)** | **<0.001** |
| Amoxicillin dispersible tablets | 89/107 (83.2) | 116/142 (81.7) | 50/56 (89.3) | 0.425 |
| Oral rehydration salts | 97/107 (90.7) | 135/142 (95.1) | 55/56 (98.2) | 0.120 |
| Ferrous sulphate plus folic acid | 81/107 (75.7) | 135/142 (95.1) | 52/56 (92.9) | <0.001 |
| Pentavalent vaccine | 5/107 (4.7) | 132/142 (93.0) | 49/56 (87.5) | <0.001 |
| Tetanus antitoxin injection | 5/107 (4.7) | 66/142 (46.5) | 28/56 (50.0) | <0.001 |
| Magnesium sulphate injection | - | 47/142 (33.1) | 48/56 (85.7) | <0.001 |
| Misoprostol 200 mcg tablets | - | 47/142 (33.1) | 34/56 (60.7) | <0.001 |
| Oxytocin injection | 3/107 (2.8) | 91/142 (64.1) | 55/56 (98.2) | <0.001 |
| **Mean availability of medicine/commodity tracer items** | **32.7 (12.8)** | **67.7 (19.5)** | **82.8 (16.3)** | **<0.001** |

Notes: Values for individual indicators are n/N (%); staff indicators denote at least one staff member. Mean tracer rows are mean facility percentage scores (SD). Denominators are all surveyed facilities of each type; skip-coded, missing, and not-applicable responses were counted as unavailable. Individual-indicator p-values are from Pearson chi-square tests; mean-score p-values are from Kruskal-Wallis tests. "Not recorded" indicates that the requested variable was absent from the supplied cleaned dataset.

**S1 Fig 1.**
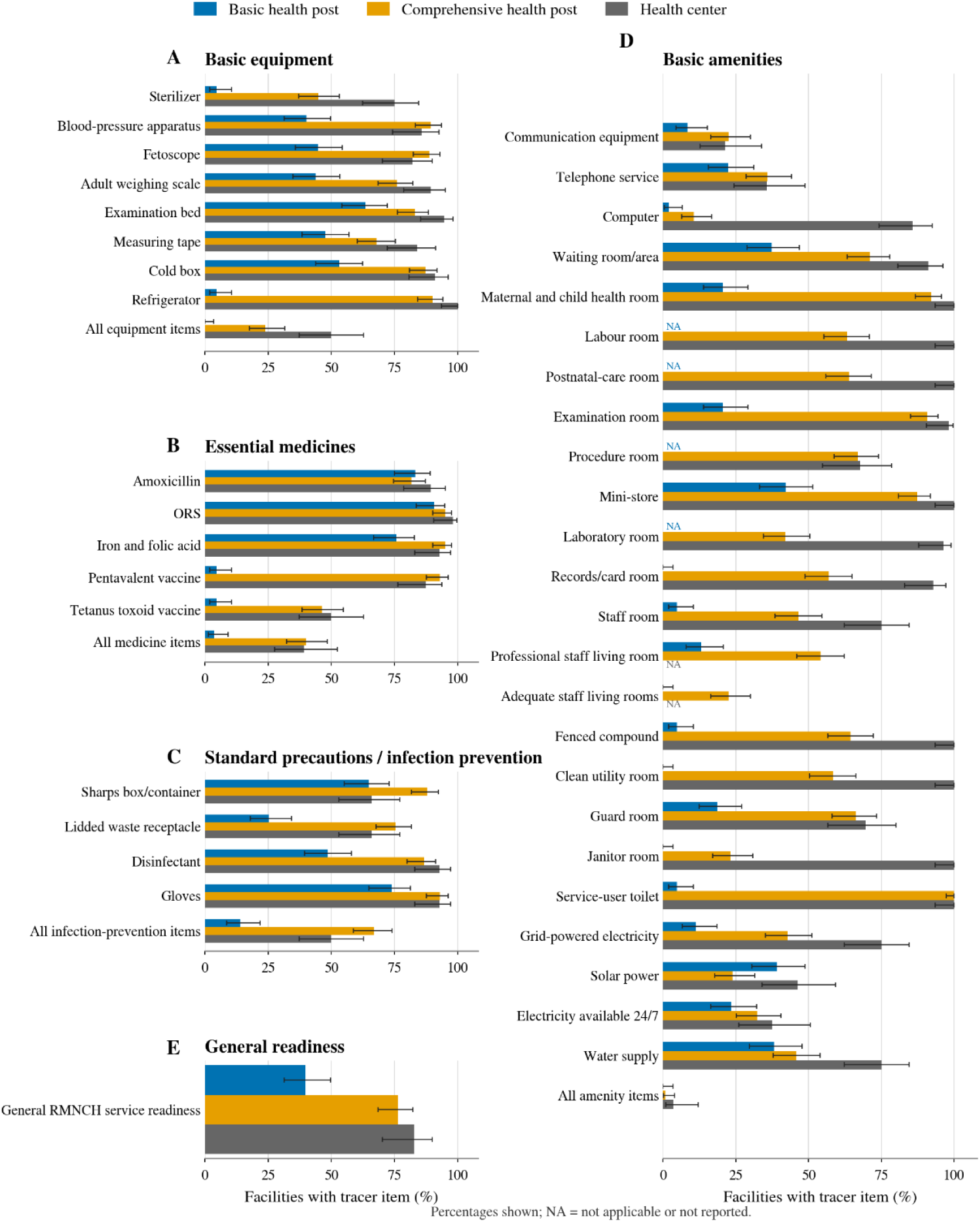
Availability of general RMNCH readiness tracer items by health-facility type.

